# Development and Evaluation of AccuPower® COVID-19 Multiplex Real-Time RT-PCR Kit and AccuPower® SARS-CoV-2 Multiplex Real-Time RT-PCR Kit for SARS-CoV-2 Detection in Sputum, NPS/OPS, Saliva and Pooled Samples

**DOI:** 10.1101/2021.11.21.21264927

**Authors:** In Bum Suh, Jaegyun Lim, Hyo Seon Kim, Guil Rhim, Kim Heebum, Kim Hana, Lee Sae-Mi, Park Hyun-sang, Song Hyun Ju, Hong MyungKook, Shin Gyung Sook, Moon Jung Kim

## Abstract

Rapid and accurate detection of the severe acute respiratory syndrome coronavirus 2 (SARS-CoV-2) is essential for the successful control of the current global COVID-19 pandemic. The real-time reverse transcription polymerase chain reaction (Real-time RT-PCR) is the most widely used detection technique. This research describes the development of two novel multiplex real-time RT-PCR kits, *AccuPower*^®^ COVID-19 Multiplex Real-Time RT-PCR Kit (NCVM) specifically designed for use with the *ExiStation*™48 system (comprised of *ExiPrep*™48 Dx and *Exicycler*™96 by BIONEER, Korea) for sample RNA extraction and PCR detection, and *AccuPower*^®^ SARS-CoV-2 Multiplex Real-Time RT-PCR Kit (SCVM) designed to be compatible with manufacturers’ on-market PCR instruments. The limit of detection (LoD) of NCVM was 120 copies/μL and the LoD of the SCVM was 2 copies/mL for both the gene and the SARS-CoV-2 gene (N gene and RdRp gene). The *AccuPower*^®^ kits demonstrated high precision with no cross reactivity to other respiratory-related microorganisms. The clinical performance of *AccuPower*^®^ kits was evaluated using the following clinical samples: sputum and nasopharyngeal/oropharyngeal swab (NPS/OPS) samples. Overall agreement of the *AccuPower*^®^ kits with a Food and Drug Administration (FDA) approved emergency use authorized commercial kit (STANDARD™ M nCoV Real-Time Detection kit, SD BIOSENSOR, Korea) was above 95% (Cohen’s kappa coefficient ≥ 0.95), with a sensitivity of over 95%. The NPS/OPS specimen pooling experiment was conducted to verify the usability of *AccuPower*^®^ kits on pooled samples and the results showed greater than 90% agreement with individual NPS/OPS samples. The clinical performance of *AccuPower*^®^ kits with saliva samples was also compared with NPS/OPS samples and demonstrated over 95% agreement (Cohen’s kappa coefficient > 0.95). This study shows the BIONEER NCVM and SCVM assays are comparable with the current standard confirmation assay and are suitable for effective clinical management and control of SARS-CoV-2.

## Introduction

Coronavirus disease 2019 (COVID-19) was first detected in Wuhan, China in 2019, and its outbreak has spread to other countries which led to a global pandemic (1). The virus that causes COVID-19 was named severe acute respiratory syndrome coronavirus 2 (SARS-CoV-2), which is the seventh known coronavirus that can infect humans (2). According to World Health Organization (WHO), as of June 30, 2021, approximately 181 million people were confirmed with SARS-CoV-2 infection, and 3.9 million were dead world-wide. (https://covid19.who.int/).

The key strategy for controlling outbreaks of COVID-19 is early and accurate detection of SARS-CoV-2 in the community. Real-time reverse transcription polymerase chain reaction (Real-time RT-PCR) a gold standard method in the detection of various viral diseases, is also the most reliable and accessible method for the diagnosis of SARS-CoV-2 infection (3). SARS-CoV-2 is a positive-sense single-strand RNA virus that consists of RNA-dependent RNA polymerase (RdRp) in an ORF1ab (4), envelope, nucleocapsid, spike, and membrane protein. The genes of these regions have been chosen as the target for the detection of SARS-CoV-2 (5). RNA viruses have a high tendency for multiple mutations. The mutations in the RNA sequence can decrease the detection ratio of the primers and probes, which may lead to the increased false-negative rates. Thus, the primers and probes in this study are designed to target multiple conserved regions to minimize the false negatives caused by mutations.

Two novel multiplex Real-Time RT-PCR kits, *AccuPower*^®^ COVID-19 Multiplex Real-Time RT-PCR Kit (Cat No. NCVM-1111, BIONEER, Korea) and *AccuPower*^®^ SARS-CoV-2 Multiplex Real-Time RT-PCR Kit (Cat No. SCVM-2112, BIONEER, Korea), were used in this study to detect the three viral genes of SARS-CoV-2 (RdRp gene, E gene, and N gene). The NCVM is a premixed product that is specifically designed to be used with the *ExiStation*™48 system (*ExiPrep*™48 Dx & *Exicycler*™96, BIONEER, Korea), in which the test is processed automatically from RNA extraction to PCR detection and confirmation. The SCVM has been developed for use with various other manufacturers’ PCR instruments. The limit of detection (LoD), cross-reactivity, and precision of the *AccuPower*^®^ kits (SCVM and NCVM) were evaluated with SARS-CoV-2 positive materials as well as commonly used human clinical samples (sputum samples and nasopharyngeal swab (NPS)/oropharyngeal swab (OPS) samples). In addition, the clinical performance of the *AccuPower*^®^ kits was verified for use in the NPS/OPS specimen pooling test and with saliva samples.

## Materials and Methods

### Primers and Probe Design

Primers and probes were designed to detect SARS-CoV-2 RNA according to two guidelines, the WHO Interim guideline and the KDCA (Korea Disease Control and Prevention Agency) guideline. The primers and probes target three different genes of SARS-CoV-2 (RdRP gene, N gene, and E gene). *In silico* analysis for inclusivity was conducted by comparing primers and probes for an alignment with all COVID-19 sequences (n=3037) in the GISAID database as of April 9th, 2020. The MUSCLE alignment was generated by multiple sequence alignment and viewed in Jalview. *In silico* analyses were performed against the updated standard database (n= 1,060,413 May 31, 2021) of the National Center for Biotechnology Information to confirm the current coverage of primers and probes. The coverage change of primers and probes was not significant. The target genes and coverage of each primer or probe are stated in Table 1.

**Table 1.** The target genes and coverage of primer and probe of the *AccuPower*® kits.

| Target Gene | Oligomer | Coverage (%) | Max. Coverage (%) | Current Coverage (%) |
| --- | --- | --- | --- | --- |
| E gene 1 | Forward primer | 99.97% | 100% | 99.94% |
|  | Reverse primer | 99.96% |  |  |
|  | Probe | 99.96% |  |  |
| E gene2 | Forward primer | 99.99% | 100% | 99.94% |
|  | Reverse Primer1 | 99.93% |  |  |
|  | Reverse Primer2 | 100% |  |  |
|  | Probe | 100% |  |  |
| RdRp gene 2-2 | Forward primer | 99.99% | 100% | 99.89% |
|  | Reverse primer | 99.98% |  |  |
|  | Probe | 99.98% |  |  |
| N gene | Forward primer | 96.67% | 99.97% | 95.06% |
|  | Reverse primer | 99.40% |  |  |
|  | Probe | 99.89% |  |  |

### *AccuPower*^®^ Kits

*AccuPower*^®^ kits contain specific primer, specific dual-labeled fluorogenic (TaqMan^®^) probe, DNA polymerase, reverse transcriptase, dNTPs, and stabilizer. Primers and fluorescent probes attach specific sequences, which distinctively appear in the SARS-CoV-2 gene. TaqMan^®^ probe contains the fluorescence in the 5’ end and the quencher in the 3’ end so the fluorescence is not released in the usual state. The fluorescence signal is emitted as the 5’-3’ exonuclease in DNA polymerase detaches from the probe, while the fluorescence and the quencher detach during PCR.

The RNA presence can be detected by fluorescence signals. The NCVM is a freeze-dried premixed product for use only with the *ExiStation*™48 system (*ExiPrep*™48 Dx & *Exicycler*™96). The SCVM is a master-mix product, which can be used with various PCR instruments. PCR reaction of *AccuPower*^®^ kits was conducted according to each manufacturer’s protocol.

### Analytical Performance Evaluation

The LoD, cross-reactivity, and precision were analyzed using the AccuPlex™ SARS-CoV-2 Verification Panel (Virus-Like Particles, SeraCare, USA) for the NCVM, and SARS-Related Coronavirus 2 (Isolate USA-WA1/2020) for the SCVM. The RNA of the SARS-CoV-2 Panel was extracted after dilution with E gene and SARS-CoV-2 gene negative NPS/OPS or sputum matrix, then the RT-PCR process was performed. RNA extraction and PCR were performed on the *ExiStation*™ 48 system. RNA extraction of the SARS-Related Coronavirus 2 was performed using the *ExiPrep*™ 48 DX (BIONEER, Korea) after dilution of the NPS/OPS or sputum matrix, following the manufacturer’s instructions.

### Limit of Detection

The LoD measurement was performed, following the CLSI guideline EP17-A2 (6). The LoD for the E gene and the SARS-CoV-2 gene were determined by measuring the RNA level in the AccuPlex™ SARS-CoV-2 Verification Panel (Virus-Like Particles, SeraCare, USA) using the NCVM, and the SARS-Related Coronavirus 2 (Isolate USA-WA1/2020) using the SCVM.

The AccuPlex™ SARS-CoV-2 Verification Panel were serially diluted to 6 concentration levels (240, 200, 160, 120, 80, 40 copies/mL) for the NCVM testing. Each dilution was tested in total 40 replicates, 20 replicates per lot, 2 lots. The SARS-Related Coronavirus 2 (Isolate USA-WA1/2020) also were serially diluted to 6 concentration levels (6, 5, 4, 3, 2, 1 copies/μL) for the SCVM testing. Each dilution was tested in total 60 replicates, 20 replicates per lot, 3 lots. Both LoD tests were performed on the *Exicycler*™96 (BIONEER, Korea) and determined using hit-rate analysis as the concentration at the lowest dilution that can be detected with >95% probability.

### Cross-reactivity

The cross-reactivity test was performed according to WHO EUL guidance (7) and Food and Drug Administration (FDA) guideline (8). The specificity of *AccuPower*^®^ kits for SARS-CoV-2 detection was evaluated by *in silico* analysis followed by testing the SARS-CoV-2 control with respiratory disease-related viruses and bacteria genes (concentration above 1.0×10^6^ copies/mL). Altogether, 29 respiratory disease-related microorganisms were tested using the NCVM and 38 were tested using the SCVM. The cross-reactivity test was performed using the *Exicycler*™96.

### Precision

The repeatability was tested according to the CLSI Guideline EP05-A3 (9) and EP15-A3 (10). The repeatability of the *AccuPower*^*®*^ kits performance was evaluated by intra-assay and inter-assay variations at different concentrations of NPS/OPS, sputum, and saliva samples.

### Ethics Statement and Sample Collection

Myongji Hospital Institutional Review Board (IRB) approved the use of surplus clinical samples for the NPS/OPS specimen-pooling (IRB No.: MJH 2020-12-028) and clinical performance evaluation (IRB No.: MJH 2020-12-029) tests. Kangwon National University Hospital IRB approved the use of saliva and NPS/OPS samples for the clinical performance testing of the *AccuPower*^®^ kits (IRB No.: KNUH-2021-03-014), which were either surplus samples or de-identified patient samples drawn after informed consent (Fig 1). RNA in the NPS/OPS and sputum samples was extracted using the *ExiPrep*™48 Viral DNA/RNA Kit (BIONEER, Korea) with the *ExiPrep*™48 Dx instrument and the RNA of saliva samples was extracted using the *ExiPrep*™48 Fast Viral RNA Kit (BIONEER, Korea) with the *ExiPrep*™48 Dx instrument. Previously, it was determined whether each sample was SARS-CoV-2 positive or negative by the confirmation test.

**Fig 1.**
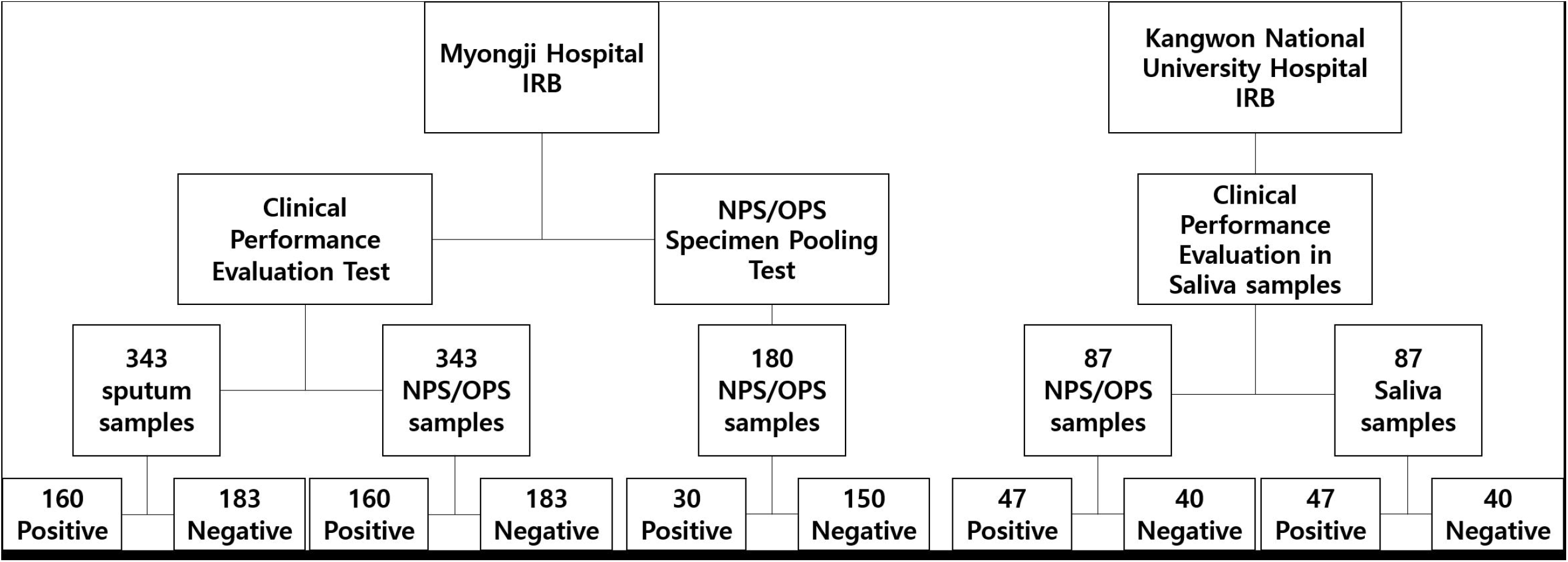
Summary of clinical specimens analyzed in this study. NPS, Nasopharyngeal Swab; OPS, Oropharyngeal Swab.

### Clinical Performance Evaluation Test

The clinical performance of *AccuPower*^®^ kits was evaluated by comparing the PCR result of each *AccuPower*^®^ kit to that of the confirmation test with confirmed positive or negative samples. A total of 343 sputum samples and 343 NPS/OPS samples were collected for clinical performance evaluation. The distribution of the Ct value of positive samples was described in Supplementary S1 Fig. At least 30 % of positive samples had Ct values within the cut-off Ct value - 10. The confirmation test was performed with the STANDARD™ M nCoV Real-Time Detection kit, the Q-Sens^®^ COVID-19 Detection Kit V2 (CancerRop, Korea), or the Allplex™ 2019-nCoV Assay (Seegene, Korea) beforehand at the sample collection institute. Presuming the result of the confirmation test was true, the clinical sensitivity and specificity of *AccuPower*^®^ kits were calculated. In addition, to validate the application of *AccuPower*^®^ kits as alternative diagnostic kits for SARS-CoV-2, the detection rate of confirmed positive and negative samples was compared to samples tested using the *AccuPower*^®^ kits and the reference kit (STANDARD™ M nCoV Real-Time Detection kit). STANDARD™ M nCoV Real-Time Detection kit is approved by the WHO and the FDA for SARS-CoV-2 detection for Emergency Use Authorization (EUA) and officially approved the first kit by the Ministry of Food and Drug Safety (MFDS), Korea. The RNA in samples was extracted using the *ExiPrep*™48 Viral DNA/RNA Kit with the *ExiPrep*™48 Dx instrument. PCR reaction was conducted using the *Exicycler*™96.

### Nasopharyngeal Swab and Oropharyngeal Swab Specimen Pooling Test

A 5-pool test on NPS/OPS samples was performed to evaluate the performance of the *AccuPower*^®^ kits on the pooled sample. A total of 180 samples (30 positive samples and 150 negative samples) were tested individually and in pools of 5 samples with the reference kit (STANDARD™ M nCoV Real-Time Detection kit) and the *AccuPower*^®^ kits. At least 25 % of the positive samples had Ct values within the cut-off Ct range of 2∼3. The 30 positive pooled samples and 30 negative pooled samples were prepared. Experimental positive pools were created using 80 µL from one SARS-CoV-2 positive specimen mixed with 4 negative patient specimens (80 µL each) for a total volume of 400 µL. Experimental negative pools included 5 negative patient specimens (80 µL each). The RNA in pooled samples was extracted using *ExiPrep*™48 Viral DNA/RNA Kit with the *ExiPrep*™48 Dx instrument. PCR reaction was conducted using the *Exicycler*™96.

The *AccuLoader*^™^ (BIONEER, Korea) which was designed to automatically load samples into a reaction well was used to pool NPS/OPS specimens to reduce human error and carry-over/cross-over contamination (Fig 2). A tablet PC connected to the *AccuLoader*^™^ controls the instrument, which consists of a barcode reader, a contamination shield cover, and a contamination prevention filter. The user inputs the sample loading positions and volumes into the PC, then, and after reading the 1D barcode attached to each sample collection tube, the instrument directs the well-plate to automatically move to the proper position which prevents sample from being loaded in the wrong well. A contamination shield cover protects other wells from splashes that may occur during sample loading and a contamination prevention filter minimizes cross-contamination from tip contact. For the pooling test, users input the number of samples and the pooled volume, and the software calculated the required volume of each sample in the pool.

**Fig 2.**
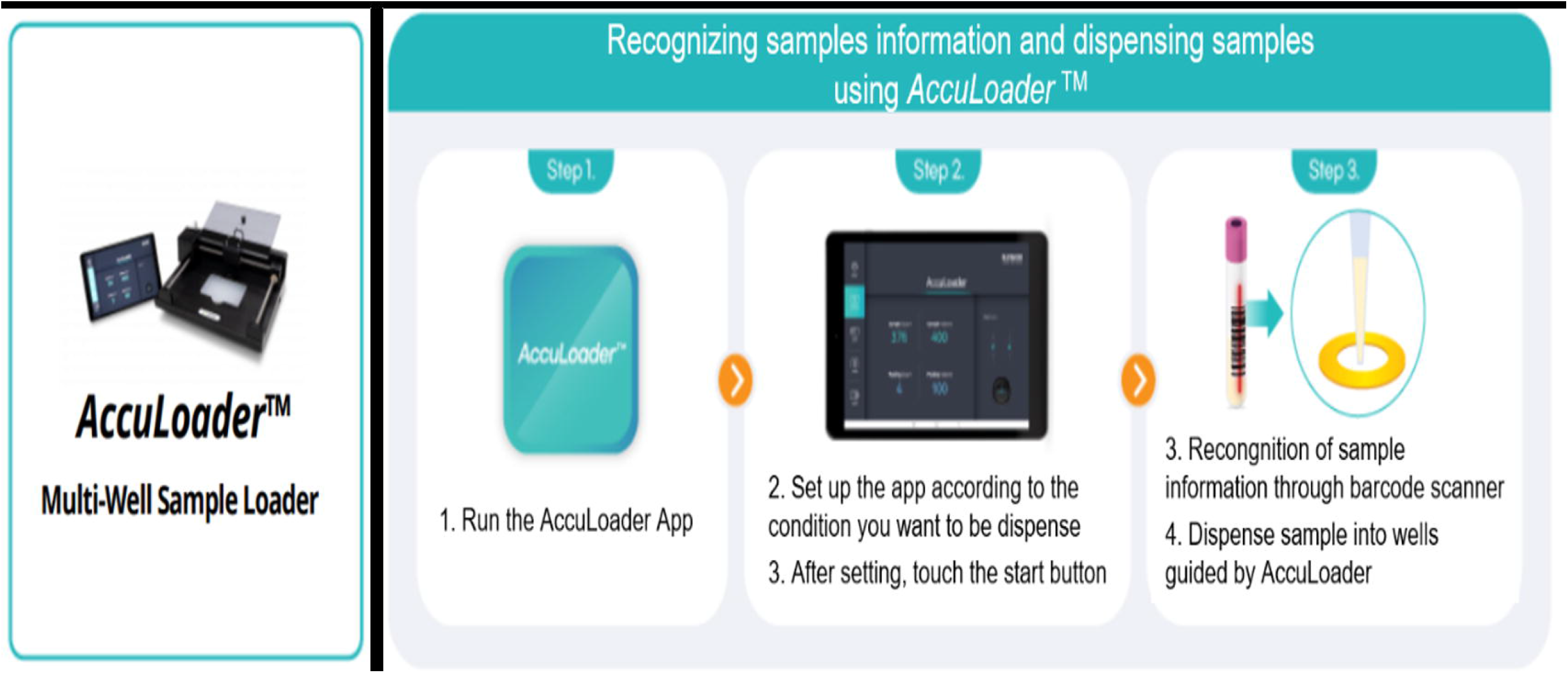
NPS/OPS specimens pooling with *the AccuLoader*™. *AccuLoader*^™^ recognizes the information of each sample by scanning the barcode on a sample tube, then guides the user to dispense samples in the correct well.

### Clinical Performance of *AccuPower*^®^ kits with Saliva Samples

The clinical performance of *AccuPower*^®^ kits with saliva samples was evaluated by analyzing the correlation between the PCR results of saliva samples and that of NPS/OPS samples. Saliva and NPS/OPS samples were collected in pairs from each patient. Patients were in a variety of stages in COVID-19 from asymptomatic period to 22 days after symptom onset. Altogether, 47 positive and 40 negative saliva and NPS/OPS paired samples were collected and stored. The RNA was extracted using the *ExiPrep*^™^48 Fast Viral RNA Kit with the *ExiPrep*^™^48 DX instrument. PCR reaction was conducted using the *Exicycler*™96.

Saliva samples were collected and stored using the Saliva Collection Kit (BIONEER, Korea), which was developed to collect, transport, and preserve saliva specimens for extraction of human genomic DNA, bacterial genomic DNA, and viral DNA/RNA for disease detection. Collection kits were gently inverted 5 times after saliva collection to properly mix the saliva and the preservation buffer.

### Statistical Analysis

Statistical analyses were conducted with R Studio 1.3.1093. In addition, the 2×2 contingency-table method was used for analyzing sensitivity, specificity, agreement, and Cohen’s kappa coefficient. Kendall’s W test was used for analyzing the correlation between PCR results in saliva samples and those in NPS/OPS samples.

## Results

### Limit of Detection

The LoD of NCVM was 120 copies/mL for the E gene (hit rate = 95%) and 120 copies/mL for the SARS-CoV-2 gene (hit rate = 95%) using the *ExiStation*^™^48 System (*ExiPrep*™48 Dx & *Exicycler*™96). The LoD of SCVM was 2 copies/μL for the E gene (hit rate = 95%), 2 copies/μL for the SARS-CoV-2 gene (hit rate = 95%) using the *ExiCycler*^™^96. (Table 2).

**Table 2.** Characteristic and PCR condition for LoD test of the *AccuPower*® kits.

| RT-PCR kit | Mix type | RNA extraction equipment | PCR equipment | Target genes | Limit of detection |
| --- | --- | --- | --- | --- | --- |
| *AccuPower*<sup>®</sup> COVID-19 Multiplex Real-Time RT-PCR Kit | Premix | *Existation*<sup>TM</sup> 48 | *Existation*<sup>TM</sup> 48 | E gene<br>*SARS-CoV-2 gene | E gene: 120 copies/mL<br>SARS-CoV-2 gene: 120 copies/mL |
| *AccuPower*<sup>®</sup> SARS-CoV-2 Multiplex Real-Time RT-PCR Kit | Master mix | *Exiprep*<sup>TM</sup> 48 DX | *Exicycler*<sup>TM</sup> 96 | E gene<br>*SARS-CoV-2 gene | E gene: 2 copies/μL<br>SARS-CoV-2 gene: 2 copies/μL |

\*SARS-CoV-2 gene: RdRp gene and N gene

### Cross-reactivity

Based on *in silico* analysis, the designed primer and probe sequences were not expected to have a significant PCR amplification by other respiratory disease-related microorganisms, and this proved to be correct. The *AccuPower*^®^ kits showed no positivity to respiratory disease-related viruses or bacteria except E gene positivity on NATrol Coronavirus-SARS Stock (qualitative, NATSARS-ST / 2003-00592). Since NATrol Coronavirus-SARS Stock originally contains the E gene, it was concluded that none of the *AccuPower*^®^ kits exhibited cross-reactions with other respiratory viruses or bacteria (Table 3).

**Table 3.** List of pathogens tested for cross-reactivity.

| No | Organism | No | Organism |
| --- | --- | --- | --- |
| 1 | Human Influenza virus A H3N2 | 20 | Parainfluenza virus 4a |
| 2 | Human Influenza virus A H1N1 | 21 | Chlamydia pneumoniae |
| 3 | Human Influenza virus B (Texas/6/11)<br>(Victoria) | 22 | Haemophilus influenzae |
| 4 | Human Coronavirus 229E | 23 | Legionella pneumophila |
| 5 | Human Coronavirus NL63 | 24 | Streptococcus pneumoniae |
| 6 | Human Coronavirus OC43 | 25 | Streptococcus pyogenes |
| 7 | Human Respiratory syncytial virus A | 26 | Bordetella pertussis |
| 8 | Human Respiratory syncytial virus B | 27 | Mycoplasma pneumoniae |
| 9 | Human Rhinovirus 14 (type B) | 28 | Pooled Human nasal wash - to represent diverse<br>microbial flora in the human respiratory tract |
| 10 | Human Metapneumovirus(hMPV) | 29 | NATrol Coronavirus-SARS Stock (qualitative)<br>(NATSARS-ST/ 2003-00592) |
| 11 | Human Adenovirus type 3 (type B) | 30 | Enterovirus 70 |
| 12 | Enterovirus 71 | 31 | Coxsackievirus B5 |
| 13 | Mycobacterium tuberculosis H37Rv | 32 | Echovirus 25 |
| 14 | MERS-CoV | 33 | Human Parachovirus 3 |
| 15 | Human coronavirus HKU1<br>(HCoV-HKU1) | 34 | Mycobacterium fortuitum |
| 16 | Adenovirus(71) | 35 | Mycobacterium intracell |
| 17 | Parainfluenza virus 1 | 36 | Mycobacterium gordonae |
| 18 | Parainfluenza virus 2 | 37 | Mycobacterium chelonae |
| 19 | Parainfluenza virus 3 | 38 | Pneumocystis jirovecii(PJP) |

Cross reactivity was evaluated using both *in silico* analysis and by PCR test. Cross reactivity of NCVM was tested with 29 pathogens (No.1 - 29). Cross-reactivity of SVCVM was tested with 38 pathogens (No.1 - 38).

### Precision

The repeatability of the *AccuPower*^®^ kits was analyzed by comparing Ct values of the PCR result within runs, between runs, and between days in each matrix at each concentration. The standard deviation (SD) of Ct values in each condition was calculated. The SD result indicates no significant difference in Ct values for positive controls within-run, between-run, between-day, and total precision. (S1 Table).

### Clinical Performance Evaluation Test

The clinical performance of the NCVM was evaluated by comparing it to the confirmation test. For sputum samples, the clinical sensitivity of NCVM was 97.50% (95% CI: 93.72% - 99.31%) and the clinical specificity was 98.36% (95% CI: 95.28% - 99.66%). For NPS/OPS samples, the clinical sensitivity the NCVM was 100.00% (95% CI: 97.72% - 100.00%) and the clinical specificity was 98.91% (95% CI: 96.11% - 99.87%) (Table 4).

**Table 4.**
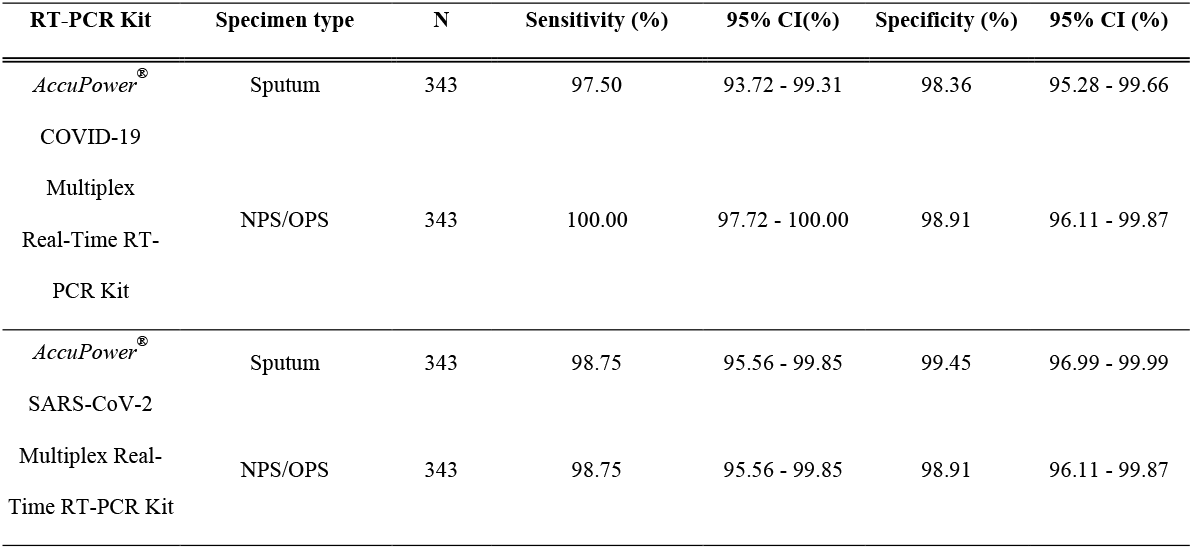
Clinical sensitivity and specificity evaluation results for the *AccuPower*® kits in Sputum or NPS/OPS specimens.

The correlation of the PCR result of the NCVM and that of the reference kit was also evaluated. For sputum samples, the positive percentage agreement was 96.27% (95% CI: 92.07% 98.62%), the negative percentage agreement was 98.90% (95% CI: 96.07% - 99.87%), the total percentage agreement was 97.38% (95% CI: 95.08% - 98.79%), and the Cohen’s kappa coefficient was 0.95. For NPS/OPS samples, the positive percentage agreement was 98.14% (95% CI: 94.65% - 99.61%), the negative percentage agreement was 98.90% (95% CI: 96.07% 99.87%), the total percentage agreement was 98.25% (95% CI: 96.23% - 99.36%), and the Cohen’s kappa coefficient was 0.97 (Table 5).

**Table 5.**
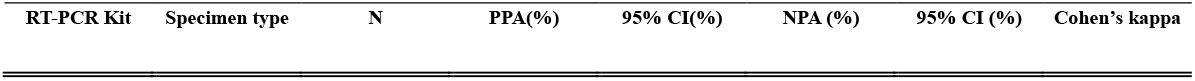
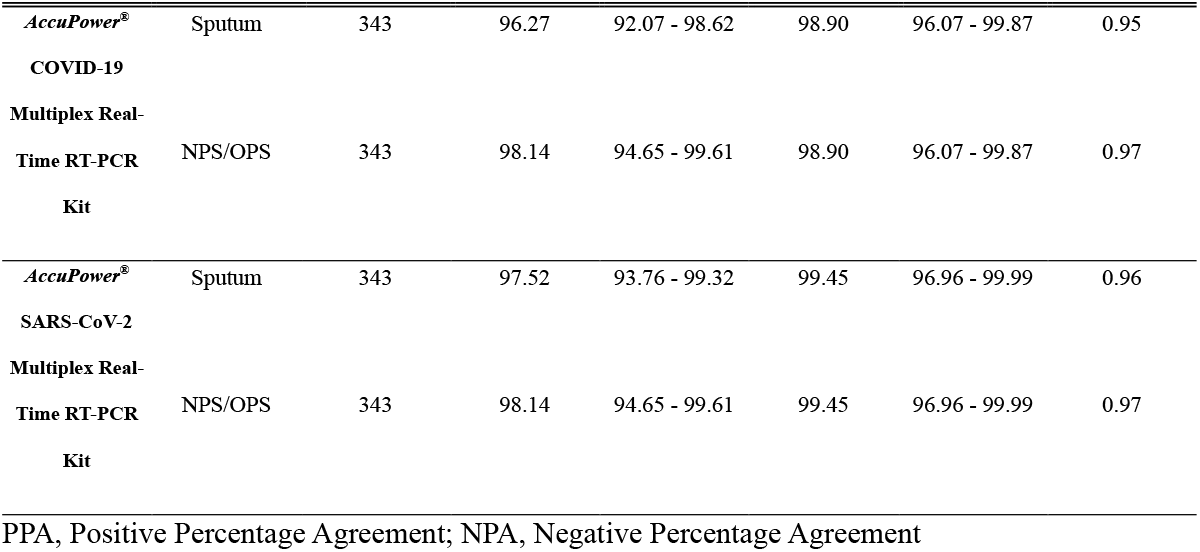
Clinical agreement evaluation results for the *AccuPower*® kits with STANDARD™ M nCoV Real-Time Detection kit.

The clinical performance of the SCVM was evaluated by comparing it to the confirmation test. For sputum samples, the clinical sensitivity of the SCVM was 98.75% (95% CI: 95.56% - 99.85%) and the clinical specificity was 99.45% (95% CI: 96.99% - 99.99%). For NPS/OPS samples, the clinical sensitivity of the SCVM was 98.75% (95% CI: 95.56% - 99.85%) and the clinical specificity was 98.91% (95% CI: 96.11% - 99.87%) (Table 4).

The correlation between results of the SCVM and that of the reference kit was evaluated. For sputum samples, the positive percentage agreement was 97.52% (95% CI: 93.76% - 99.32%), the negative percentage agreement was 99.45% (95% CI: 96.96% - 99.99%), the total percentage agreement was 98.25% (95% CI: 96.23% - 99.36%), and the Cohen’s kappa coefficient was 0.96. For NPS/OPS samples, the positive percentage agreement was 98.14% (95% CI: 94.65% - 99.61%), the negative percentage agreement was 99.45% (95% CI: 96.96% - 99.99%), the total percentage agreement was 98.54% (95% CI: 96.63% - 99.53%), and the Cohen’s kappa coefficient was 0.97 (Table 5).

The correlation of Ct values in target genes among NCVM, SCVM, and STANDARD^™^ M nCoV Real-Time Detection kit was analyzed by correlation analysis with the plot in R studio and showed significant correlations with coefficient of determination (R^2^) at ≥0.97 (S2 Fig). An analysis of Ct values was also performed using the ANOVA test with master mix kits (STANDARD^™^ M nCoV Real-Time Detection kit and SCVM). The ANOVA test showed no significant difference between SCVM and STANDARD^™^ M nCoV Real-Time Detection kit (P>0.05) (S3 Fig).

### Nasopharyngeal Swab and Oropharyngeal Swab Specimen Pooling Test

Pooled samples were prepared as described above (S2 Table). Samples were then tested individually using the *AccuPower*^®^ kits and the reference kit. The test showed 100% positive and negative agreement of the *AccuPower*^®^ kits with the reference kit. The clinical performance of the *AccuPower*^®^ kits was also tested using in 5-pooled samples and evaluated by comparing the PCR results from the *AccuPower*^®^ kits to that of the reference kit. For NCVM, the positive percentage agreement was 93.30% (95% CI: 77.93% - 99.18%) and the negative percentage agreement was 100.00% (95% CI: 88.43% - 100.00%) with the reference test of pooled samples.

The positive and negative agreement between pooled samples and individual samples was also analyzed. For NCVM, the positive percentage agreement of pooled samples was 93.30% (95% CI: 77.93% - 99.18%) and the negative percentage agreement was 100.00% (95% CI: 88.43% - 100.00%), compared to individual samples. The average Ct value of pooled samples was 1.65 higher than the average Ct value of individual samples in the E gene and 1.90 higher in the SARS-CoV-2 gene (Fig 3.A). The degree of association between the Ct value of individual samples and the Ct value of pooled samples was analyzed by regression analysis and expressed as y=0.887x+4.885, R^2^=0.9700 in the E gene and y=0.983x+2.375, R^2^=0.9955 in the SARS-CoV-2 gene (Fig 3.B).

**Fig 3.**
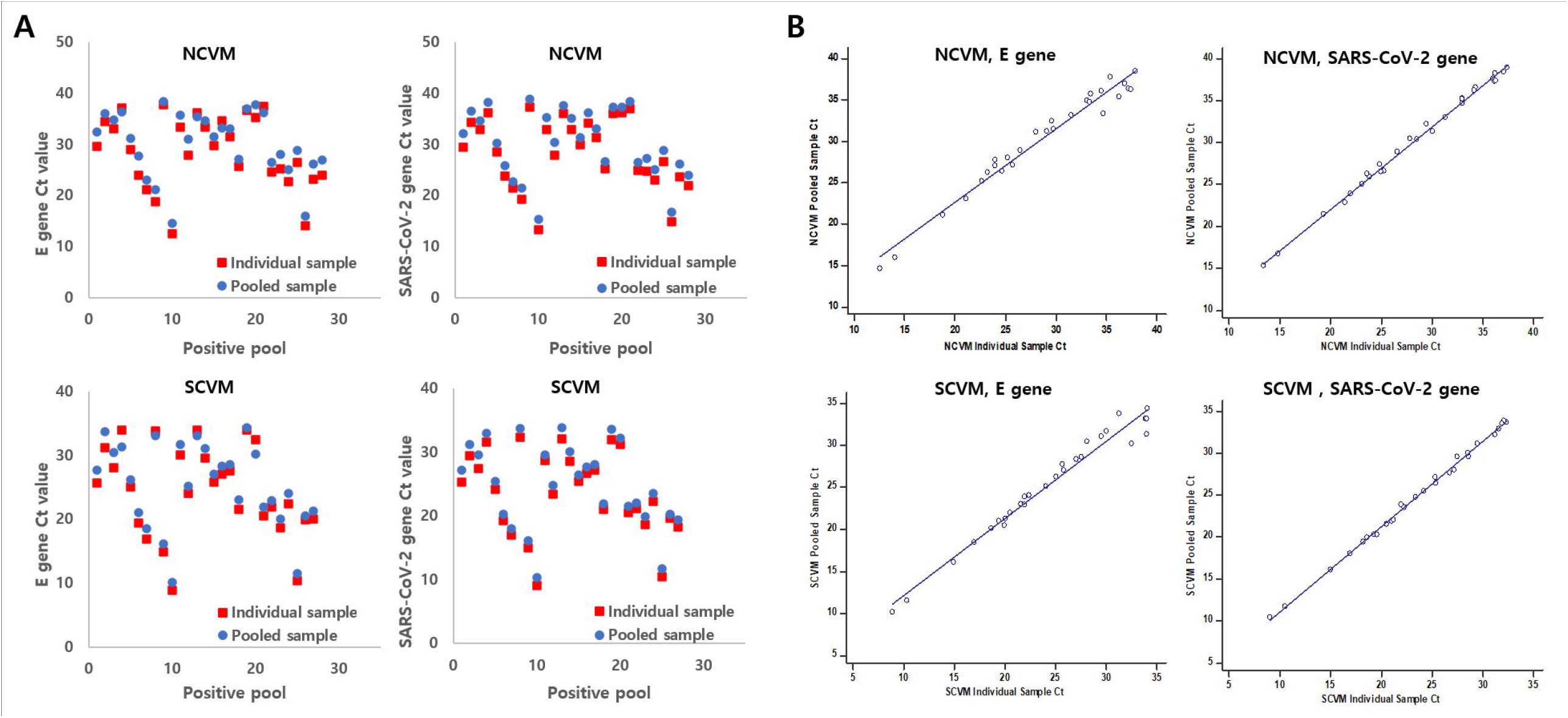
The comparison of Ct values in individual samples and pooled samples in NPS/OPS specimen pooling test. (A) Ct values of individual and pooled samples in the *AccuPower*^®^ kits. (B) The regression analysis with Ct values in individual samples and pooled samples in the *AccuPower*^®^ kits.

For SCVM, the positive percentage agreement was of pooled samples 90.00% (95% CI: 73.47% −97.89%) and the negative percentage agreement was 100.00% (95% CI: 88.43% - 100.00%) compared to the reference test. In addition, the positive percentage agreement was 90.00% (95% CI: 73.47% −97.89%) and the negative percentage agreement was 100.00% (95% CI: 88.43% - 100.00%) in pooled sample, compared to individual samples. The average Ct value of pooled samples was 0.94 higher than the average Ct value of individual samples in the E gene and 1.25 higher in the SARS-CoV-2 gene (Fig 3.A). The degree of association between the Ct value of individual samples and the Ct value of pooled samples was analyzed by regression analysis and expressed as y=0.922x+2.841, R^2^=0.9716 in E gene and y=1.017x+0.840, R^2^=0.9976 in SARS-CoV-2 gene (Fig 3.B). The results of the pooling tests are described in Table 6. The disagreement between the PCR results of pooled samples and those of individual samples has occurred only in samples with low concentration (Ct>Cutoff Ct - 3) of target genes (S3 Table).

**Table 6.** The pooling test evaluation results for the *AccuPower*® kits.

| Major discordance | Individual test (reference) |  |  |  |
| --- | --- | --- | --- | --- |
|  | No.positive | No.negative | No.positive | No.negative |
|  | Individual test (NCVM) |  | Pool test (NCVM) |  |
| Positive | 30 | 0 | 28 | 0 |
| Negative | 0 | 150 | 2 | 30 |
| PPV (%) | 100.00% (95% CI: 88.43% - 100.00%) |  | 93.30% (95% CI: 77.93% - 99.18%) |  |
| NPV (%) | 100.00% (95% CI: 97.57% - 100.00%) |  | 100.00% (95% CI: 88.43% - 100.00%) |  |
| Accuracy (%) | 100.00% |  | 96.67% |  |
| cohen's kappa ($\kappa$) | 1.00 | | 0.93 | |
|  | No.positive | No.negative | No.positive | No.negative |
|  | Individual test (SCVM) |  | Pool test (SCVM) |  |
| Positive | 30 | 0 | 27 | 0 |
| Negative | 0 | 150 | 3 | 30 |
| PPV (%) | 100.00% (95% CI: 88.43% - 100.00%) |  | 90.00% (95% CI: 73.47% - 97.89%) |  |
| NPV (%) | 100.00% (95% CI: 97.57% - 100.00%) |  | 100.00% (95% CI: 88.43% - 100.00%) |  |
| Accuracy (%) | 100.00% |  | 95.00% |  |
| cohen's kappa ($\kappa$) | 1.00 | | 0.90 | |

### Clinical Performance of *AccuPower*^®^ kits with Saliva Samples

The clinical performance of the *AccuPower*^®^ kits for use with saliva samples was evaluated by comparing the PCR results in saliva samples to those in NPS/OPS samples. The Ct values of each *AccuPower*^®^ kit using both kinds of samples from diverse stages of COVID-19 are shown in Fig 4.A.

**Fig 4.**
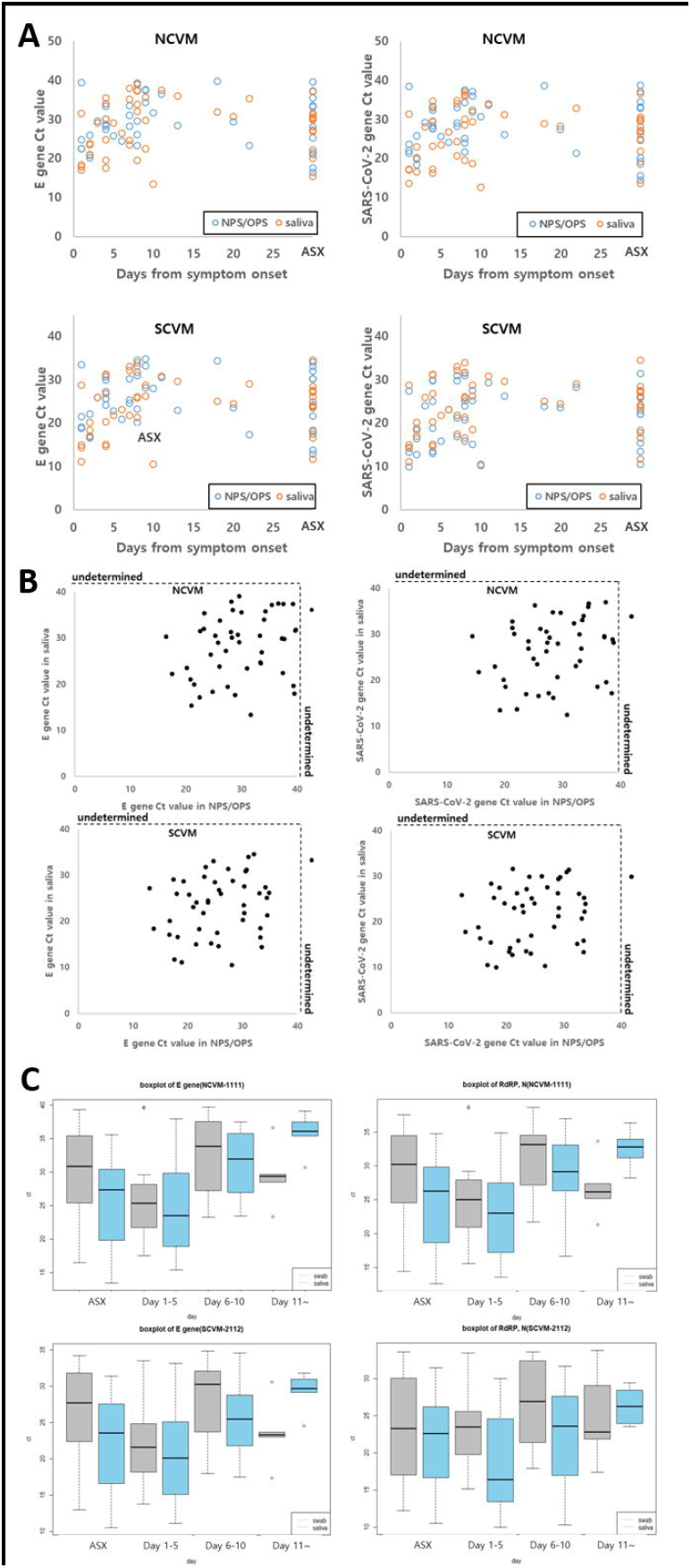
Clinical performance of the *AccuPower*® kits in saliva and NPS/OPS samples. (A) Ct values of saliva and NPS/OPS samples from patients in various stages of COVID-19. (B) Clinical performance comparison of the *AccuPower*^®^ kits in saliva and NPS/OPS samples shown in scatter plots. (C) Ct value comparison by boxplot among the NCVM, SCVM, and STANDARD^™^ M nCoV Real-Time Detection kit. ASX, Asymptomatic.

The PCR results of each *AccuPower*^®^ kit used with each type of sample were verified by comparison to the confirmation test. The NCVM PCR results for NPS/OPS samples showed 97.87% (95% CI: 88.71% - 99.95%) positive agreement, the 100.00% (95% CI: 91.19% - 100.00%) negative agreement, and 98.85% (95% CI: 93.76% - 99.97%) total agreement with a Cohen’s kappa coefficient of 0.98, compared to the PCR results of the confirmation test for NPS/OPS samples. The NCVM PCR results for saliva samples showed 100.00% (95% CI: 92.45% - 100.00%) positive agreement, 100.00% (95% CI: 91.19% - 100.00%) negative agreement, and 100.00% (95% CI: 95.85% - 100.00%) total agreement with a Cohen’s kappa coefficient of 1.00, when compared to the PCR results of the confirmation test for NPS/OPS samples. The comparison of the NCVM PCR results between NPS/OPS samples and saliva samples showed that 100.00% (95% CI: 92.29% - 100.00%) positive agreement, 97.56% (95% CI: 87.14% - 99.94%) negative agreement, and 98.85% (95% CI: 93.76% - 99.97%) total agreement with Cohen’s kappa coefficient of 0.98.

The SCVM PCR results for NPS/OPS samples showed 97.87% (95% CI: 88.71% - 99.95%) positive agreement, 100.00% (95% CI: 91.19%- 100.00%) negative agreement, and 98.85% (95% CI: 93.76% - 99.97%) total agreement with a Cohen’s kappa coefficient of 0.98, compared to the PCR results of the confirmation test for NPS/OPS samples. The SCVM PCR results for saliva samples showed 100.00% (95% CI: 92.45% - 100.00%) positive agreement, 100.00% (95% CI: 91.19% - 100.00%) negative agreement, and 100.00% (95% CI: 95.85% - 100.00%) total agreement with a Cohen’s kappa coefficient of 1.00, compared to the PCR results of the confirmation test for NPS/OPS samples. The comparison of the SCVM PCR results between NPS/OPS samples and saliva samples showed 100.00% (95% CI: 92.29% - 100.00%) positive agreement, 97.56% (95% CI: 87.14% - 99.94%) negative agreement, and 98.85% (95% CI: 93.76% - 99.97%) total agreement, with a Cohen’s kappa coefficient of 0.98. The results indicate the adequacy of the *AccuPower*^®^ kits for the use with saliva samples (Table 7).

**Table 7.**
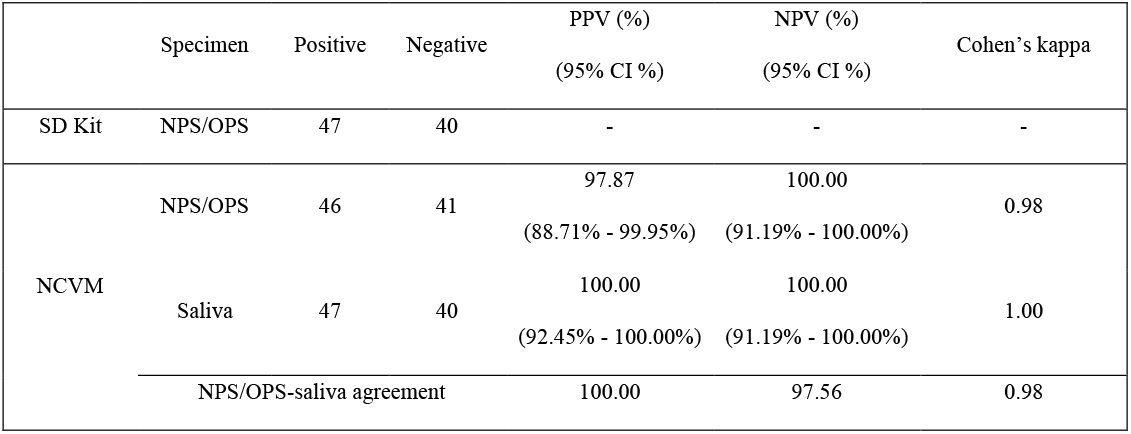
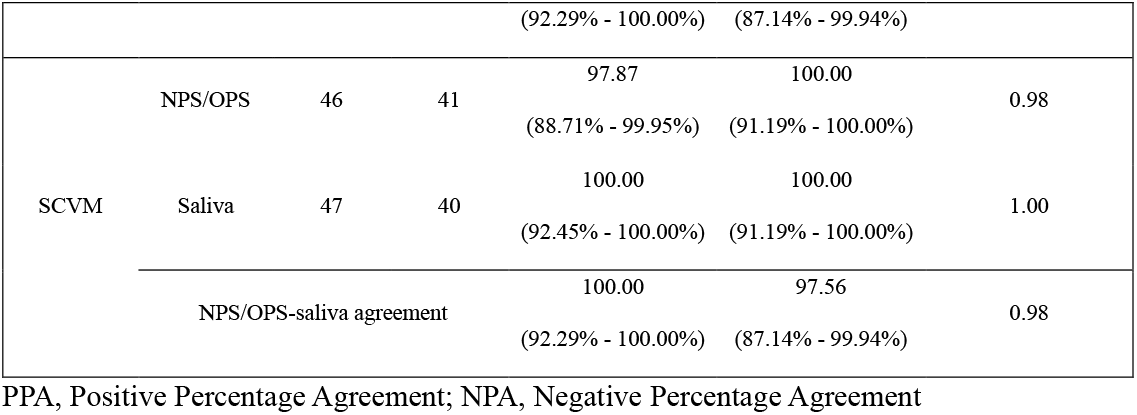
Clinical performance evaluation results of the *AccuPower*® kits in saliva samples.

The scatter plots of Ct values in paired NPS/OPS and saliva specimens were analyzed for each *AccuPower*^®^ kit (Fig 4.B). The correlation of Ct values in paired NPS/OPS and saliva specimens showed no significant difference in the E gene and the SARS-CoV-2 gene. Kendall’s W was over 0.5 in the E gene (W=0.639 for NCVM, 0.596 for SCVM) and the SARS-CoV-2 gene (W=0.613 for NCVM, 0.589 for SCVM), showing a high degree of agreement. In addition, the association of Ct values in each type of sample with days from the onset of COVID-19 was examined (Fig 4.C). Up to 10 days from the onset of COVID-19, Ct values of saliva samples were lower than those of NPS/OPS samples. On the other hand, after 10 days from the onset of COVID-19, Ct values of NPS/OPS samples were lower than those of saliva samples.

## Discussion

The COVID-19 pandemic became the catalyst for the development of more rapid and accurate detection methods for SARS-CoV-2 to better support the clinicians and front-line healthcare professionals (11). While effective vaccines have been developed, the availability of high-quality diagnostic methods remains essential (12). Many studies continue to target a more efficient, reliable, and sensitive detection method for SARS-CoV-2. On the other hand, multiplex RT-PCR viral RNA detection assays have been developed for fast and reliable SARS-CoV-2 detection (13-16).

The analytical performance of two *AccuPower*^®^ kits (NCVM as the premix type and SCVM as the master mix type) was evaluated. The NCVM was designed to be used on a closed system (*ExiStation*™48 system) to provide full automation and contamination-free and error-free results. The LoD of the NCVM was 120 copies/mL for the E gene and the SARS-CoV-2 gene as determined by the *ExiStation*™48 system. On the other hand, the SCVM was designed to be used on an open system compatible with other manufacturers’ PCR instruments. The LoD of the SCVM was 2 copies/ul for the E gene and the SARS-CoV-2 gene as determined by the *Exicycler*^™^ 96. In addition, The LoD was determined by multiple alternative PCR instruments (CFX96™ Real-Time PCR Detection System (Bio-rad, USA), Applied Biosystems 7500 Fast Real-time PCR Instrument system (Thermo Fisher Scientific, USA), QuantStudio™5 Real-Time PCR Instrument (Thermo Fisher Scientific), *Exicycler*™384 (BIONEER, Korea), *Exicycler*™96 Fast (BIONEER, Korea)) and the results demonstrated comparable performance (S4 Table). Cross-reactivity was tested with 29 respiratory disease-related viruses and bacteria genes were performed for the NCVM, and with 38 for the SCVM, respectively. The results showed that there was no detectable cross-reactivity in *AccuPower*^®^ kits, Precision of the two *AccuPower*^®^ kits was evaluated and the results showed high within-run, between-run, between-day, and total precision.

This study provides the data to support the usability of the *AccuPower*^®^ kits for the detection of SARS-CoV-2 RNA in sputum and NPS/OPS clinical samples as evidenced by equivalency with the confirmation test, which was performed in the collection institute beforehand, and with the reference kit (STANDARD™ M nCoV Real-Time Detection kit). Specifically, the results of the *AccuPower*^®^ kits were validated with over 95% agreement with both the confirmation test and the reference kit. On the other hand, the NPS/OPS specimen pooling test showed the PCR result of the pooled samples of 5 individuals had over 90% agreement with the individual samples by the *AccuPower*^®^ kits. The PCR results of the *AccuPower*^®^ kits in saliva samples showed over 95% positive and negative agreement with those in the NPS/OPS samples, confirming the usability of the *AccuPower*^®^ kits for the saliva samples as well. In addition, the correlation between Ct values and days from the onset of COVID-19 in both NPS/OPS and saliva samples indicates the saliva samples are more suitable for detection up to 10 days from the onset of COVID-19 and the NPS/OPS samples are more suitable for detection after 10 days from the onset of COVID-19 for SARS-CoV-2 detection. This result is consistent with the previous studies on the COVID-19 saliva samples (17). In brief, the analytical and clinical performance of the *AccuPower*^®^ kits showed they are as effective in the SARS-CoV-2 detection kit as the current standard confirmation test including the reference kit.

A previous study presented the LoD of *AccuPower*^®^ kits without the appropriate number of replications, using quantified specimens RNA for the test (18). On the contrary, in this study, The LoD test was performed with 20 replicates, which is recommended in the CLSI guideline, using SARS-CoV-2 verification panel and SARS-Related Coronavirus 2. In addition, the clinical performance of each *AccuPower*^®^ kit was tested with clinical samples of various Ct and verified by comparing them to the performance of STANDARD™ M nCoV Real-Time Detection kit, which obtained WHO and FDA approval for emergency use and MFDS official approval and showed high agreements (>95%). STANDARD™ M nCoV Real-Time Detection kit showed higher clinical sensitivity with Allplex™ 2019-nCoV Assay in a comparison study (15). Also, the clinical performance evaluation of *AccuPower*^®^ kits, assuming the result of Allplex™ 2019-nCoV Assay, which was one type of test in the confirmation test, as true, indicated high sensitivity and specificity (>95%) (S5 Table). Thus, it may be suggested that *AccuPower*^®^ kits have equivalent clinical sensitivity with Allplex™ 2019-nCoV Assay.

In conclusion, this study describes the successful development of two multiplex real-time RT-PCR methods, NCVM and SCVM, for the diagnosis of SARS-CoV-2. Simultaneous targeting of three viral genes (RdRP, N, and E) by the *AccuPower*^®^ kits provides an accurate, reliable, and easy-to-use SARS-CoV-2 detection test. The *AccuPower*^®^ kits demonstrate the analytical performance characteristics expected of a valid diagnostic assay. The clinical performance of the *AccuPower*^®^ kits was comparable to the gold standard confirmation test, including the reference kit. In addition, the specimen pooling test with n=5 showed the ability of the *AccuPower*^®^ to process high volume samples cost-effectively for use as a surveillance tool. The clinical performance test of *AccuPower*^®^ kits in saliva samples demonstrated the usability of the *AccuPower*^®^ kits with saliva samples and the saliva samples being more adequate than NPS/OPS samples for early detection (before 10 days from the onset of symptom) of COVID-19. The *AccuPower*^®^ assay can be used for the fast and dependable detection of the SARS-CoV-2 virus.

## Data Availability

All data produced in the present study are available upon reasonable request to the authors

## Acknowledgments

This research was supported by a grant of the Korea Health Technology R&D Project through the Korea Health Industry Development Institute (KHIDI), funded by the Ministry of Health & Welfare, Republic of Korea (grant number: HI20C2557).

## Supporting information

**S1 Fig. Ct value variation in collected clinical samples**. Nashpharyngeal and Oropharyngeal swab specimens Ct distribution (Above). Sputum specimens Ct distribution (Below). *SD: STANDARD™ M nCoV Real-Time Detection kit, Cutoff (Ct): 36 **CancerRop: Q-Sens^®^ COVID-19 Detection Kit V2, Cutoff (Ct): 40 ***Seegene: Allplex™ 2019-nCoV Assay, Cutoff (Ct): 40

**S2 Fig. Correlation Analysis with plot among NCVM, SCVM, and STANDARD™ M nCoV Real-Time Detection kit**. Ct values among assays (NCVM, SCVM and STANDARD™ M nCoV Real-Time Detection kit) showed high correlation with a Pearson R^2^ correlation coefficient ≥0.97.

**S3 Fig. Comparison between SCVM and STANDARD™ M nCoV Real-Time Detection kit**. Results of ANOVA test showed no significant difference between SCVM and STANDARD™ M nCoV Real-Time Detection kit (p>0.05).

**S1 Table. Precision evaluation results for the *AccuPower*® kits**. Precision evaluation results of *AccuPower*^®^ COVID-19 Multiplex Real-Time RT-PCR Kit and *AccuPower*^®^ SARS-CoV-2 Multiplex Real-Time RT-PCR Kit.

**S2 Table. Individual samples and pooled samples in swab specimen pooling test**.

**S3 Table. The agreement between individual samples and pooled samples in the *AccuPower*® kits**.

**S4 Table. Limit of detection of SCVM in various PCR instruments**.

**S5 Table. Clinical sensitivity and specificity evaluation results for the *AccuPower*® kits in Sputum or NPS/OPS specimens, compared to Allplex™ 2019-nCoV Assay**.

