## Supplementary figures and images for "Development and Evaluation of AccuPower® COVID-19 Multiplex Real-Time RT-PCR Kit and AccuPower® SARS-CoV-2 Multiplex Real-Time RT-PCR Kit for SARS-CoV-2 Detection in Sputum, NPS/OPS, Saliva and Pooled Samples"

### S1 Fig

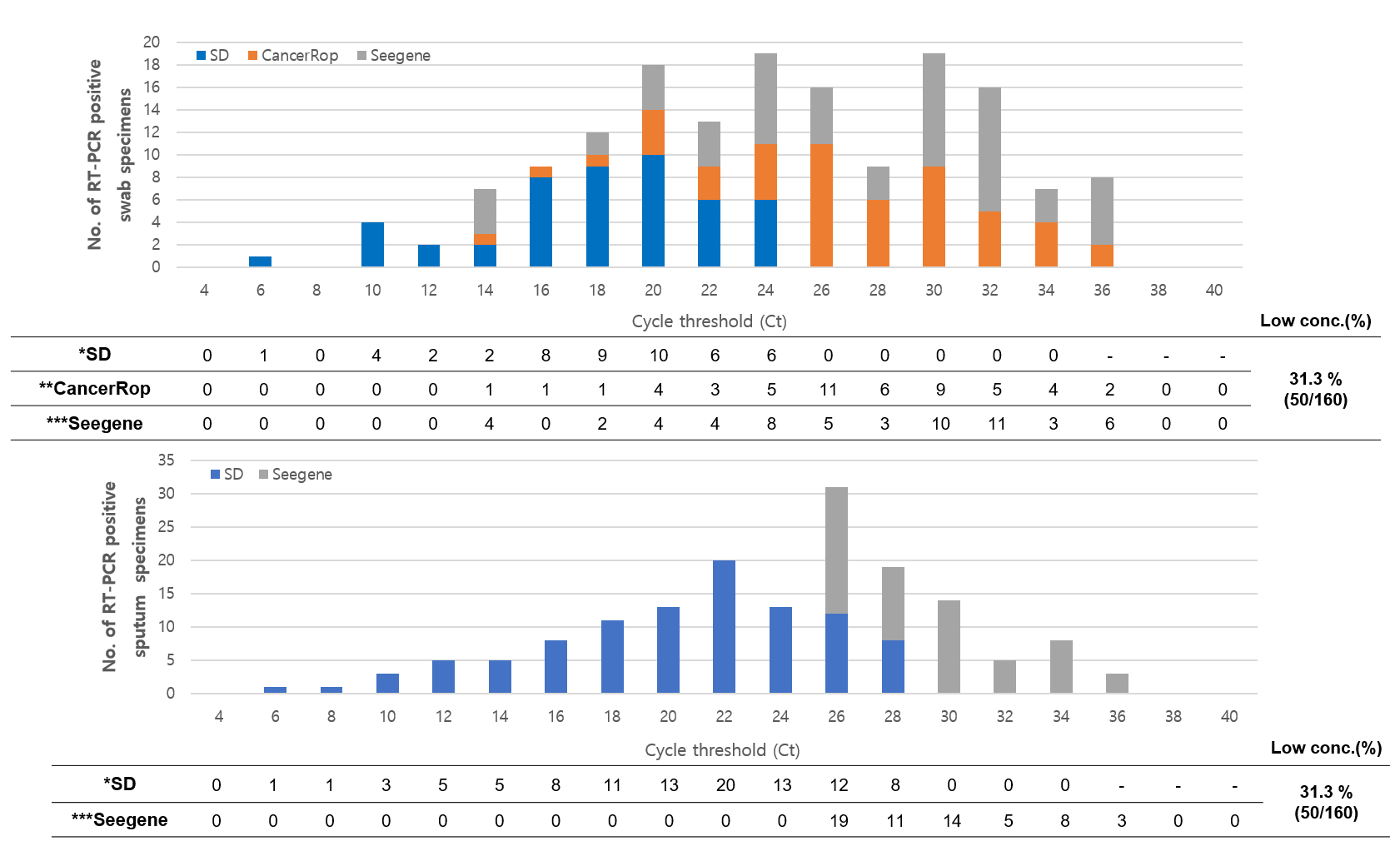

### S1 Table

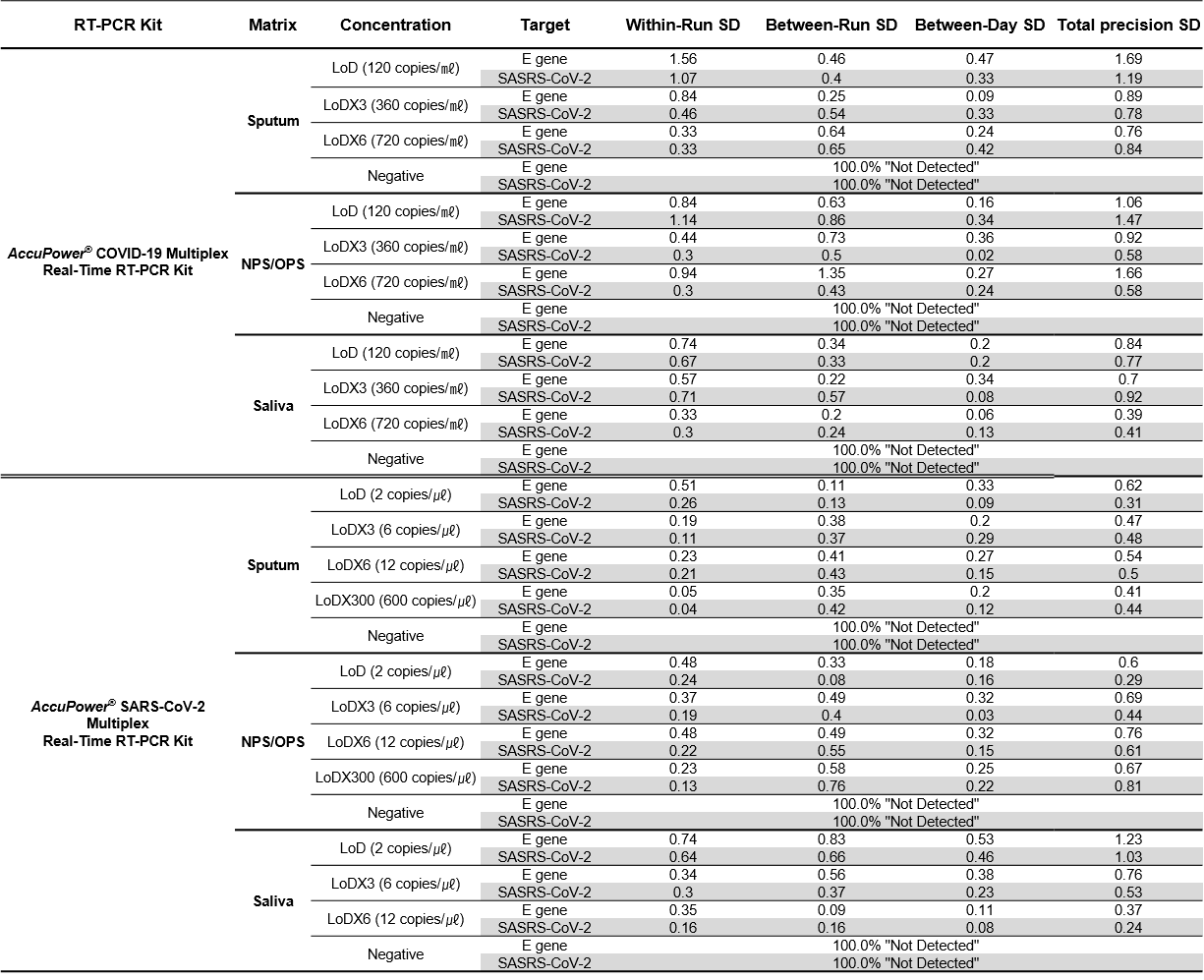

### S2 Fig

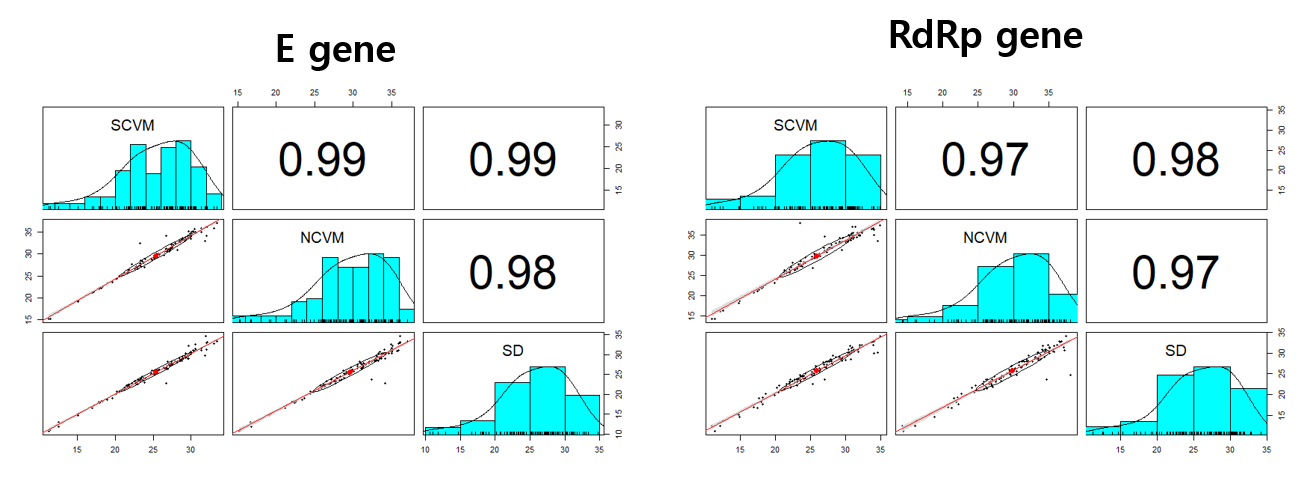

### S2 Table

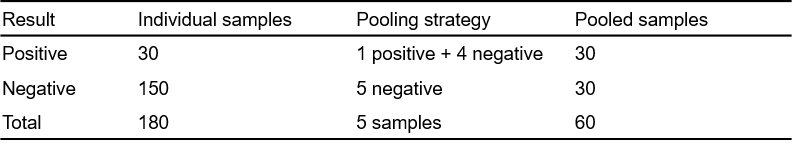

### S3 Fig

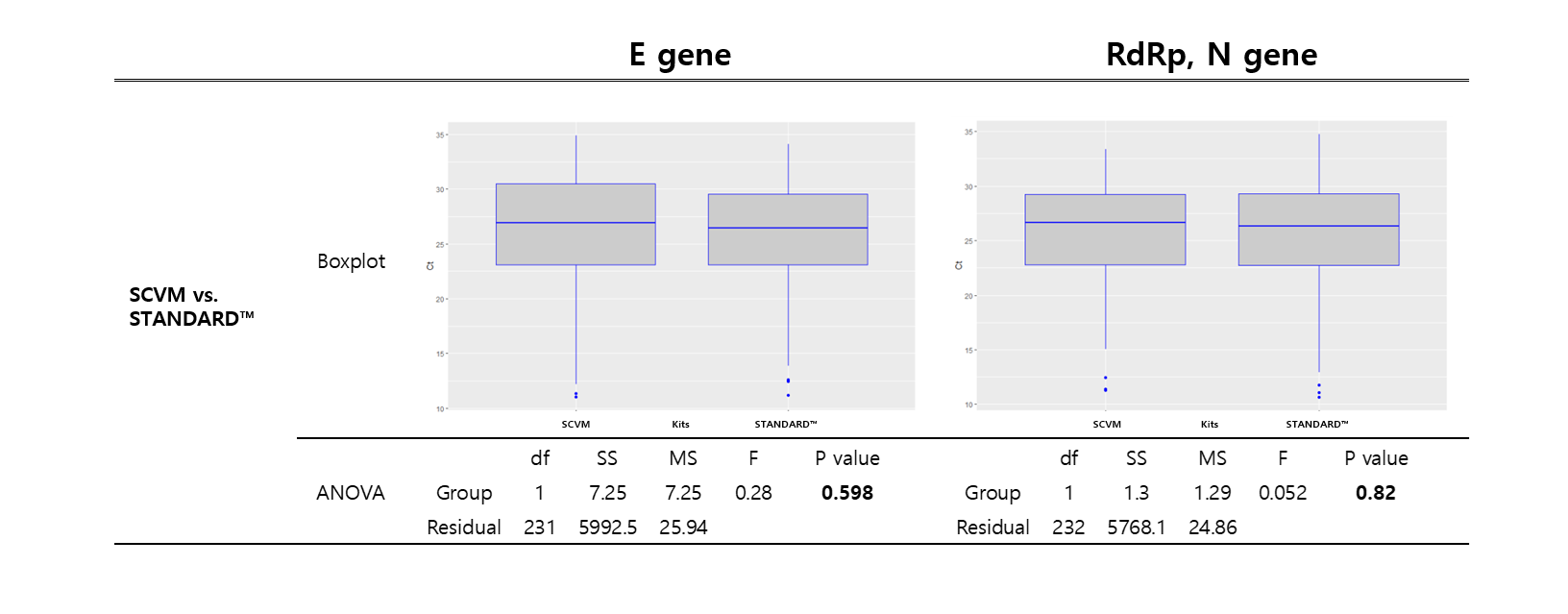

### S3 Table

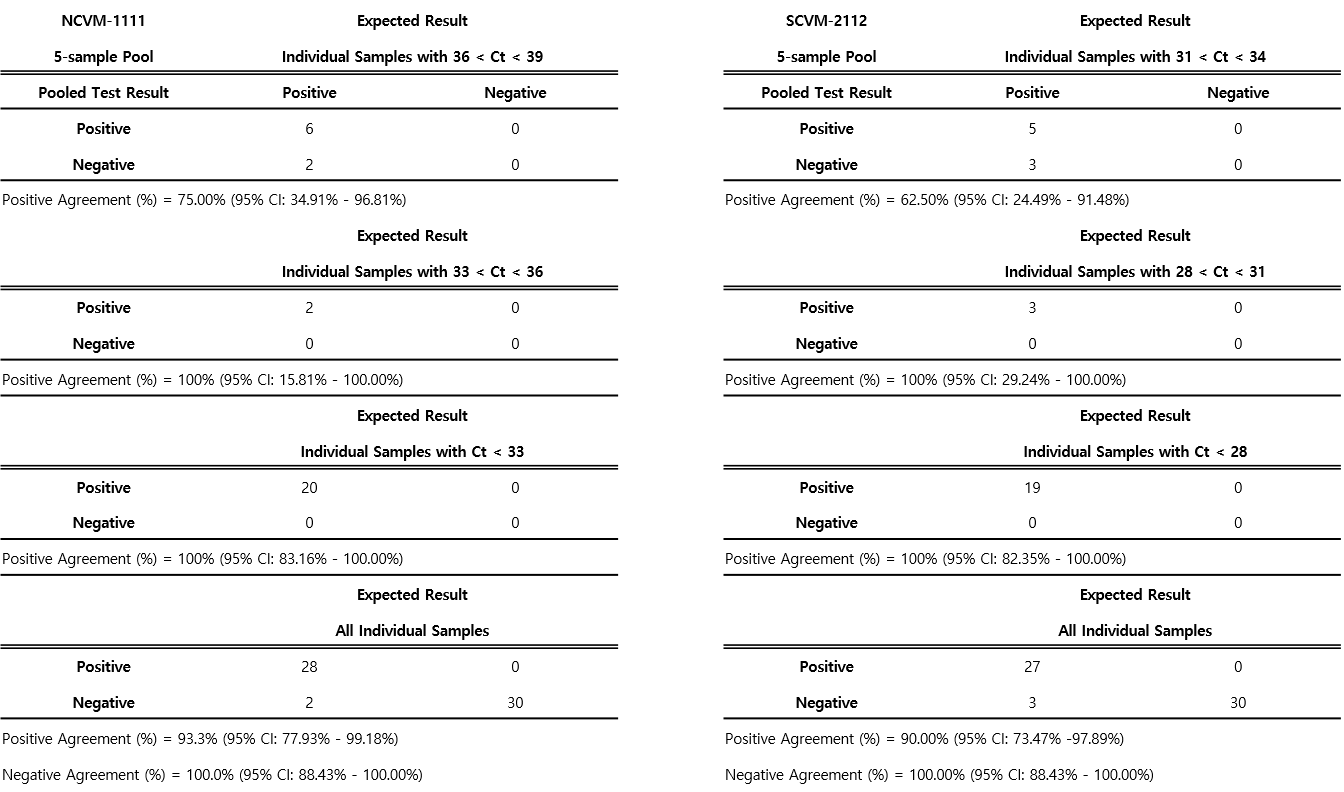

### S4 Table

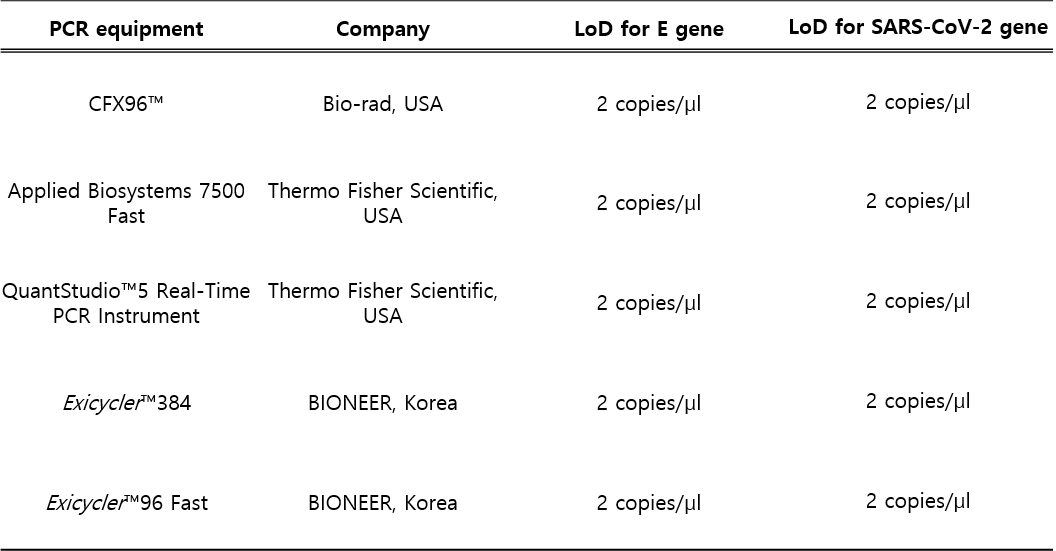

### S5 Table

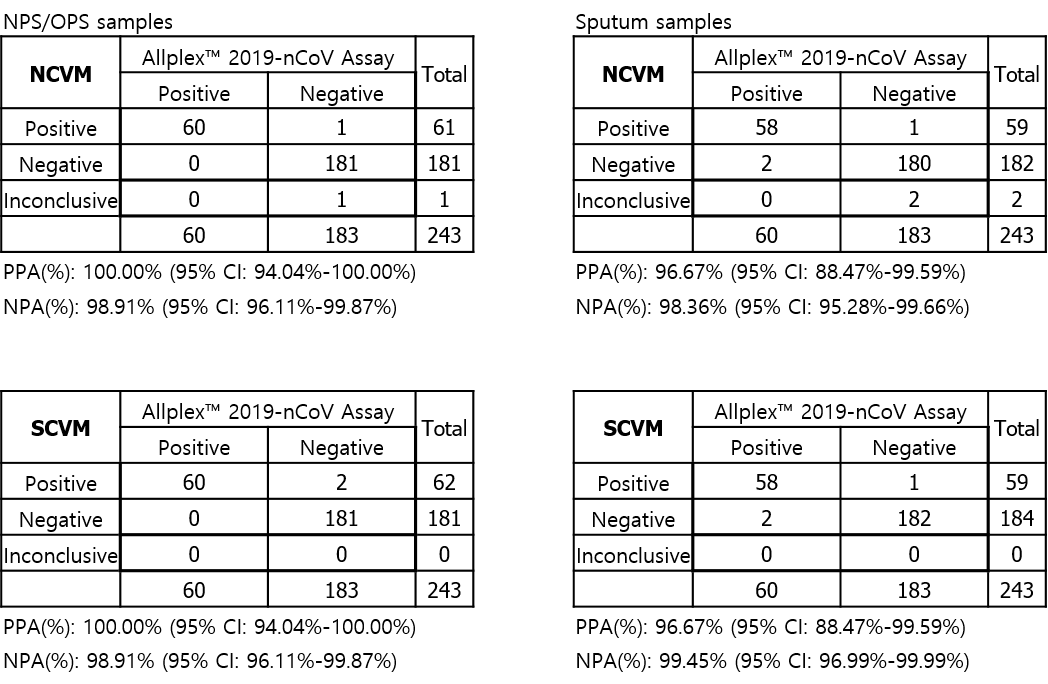
